# Hepatitis B Virus (HBV) prevalence and characteristics in HIV-transmitting mothers and their infants in KwaZulu-Natal, South Africa

**DOI:** 10.1101/2023.03.20.23287409

**Authors:** Jane Millar, Gabriela Z.L. Cromhout, Noxolo Mchunu, Nomonde Bengu, Thumbi Ndung’u, Philip J Goulder, Philippa C Matthews, Anna L McNaughton

## Abstract

**Background:** HIV and hepatitis B virus (HBV) prevalence are high in KwaZulu-Natal (KZN), South Africa. HIV co-infection negatively impacts HBV prognosis, and can increase likelihood of HBV mother-to-child-transmission (MTCT). In an established early treatment intervention cohort of HIV-transmitting mother-child pairs in KZN, we characterised HBV serological makers in mothers, and screened at-risk infants for HBV.

**Methods:** Maternal samples (n=175) were screened for HBV infection (HBsAg), exposure to HBV (anti-HBc) and vaccination responses (anti-HBs-positive without other HBV markers). Infants of HBV-positive mothers were screened for HBsAg at 1 and 12 months.

**Results:** HBV infection was present in 8.6% (15/175) of mothers. Biomarkers for HBV exposure were present in 31.4% (55/175), but absent in 53.3% (8/15) maternal HBV-positive cases. Maternal HBV vaccination appeared rare (8.0%; 14/175). Despite prescription of antiretroviral therapy (ART) active against HBV, HBV DNA was detectable in 46.7% (7/15) HBsAg-positive mothers, with (5/7) also viraemic for HIV. Three mothers had HBV viral loads >5.3log_10_ IU/ml, making them high-risk for HBV MTCT. Screening of available infant samples at one month of age (n=14) found no cases of HBV MTCT, and at 12 months (n=13) identified one HBV infection. Serological vaccination evidence was present in 53.8% (7/13) infants tested.

**Discussion:** This vulnerable cohort of HIV-transmitting mothers had a high undiagnosed HBV prevalence. Early infant ART may have reduced risk of MTCT in high-risk cases. Current HBV guidelines recommend antenatal antiviral prophylaxis but these data underline a potential role for infant post-exposure prophylaxis in high-risk MTCT pairs, warranting further investigation.

## Introduction

Approximately 316 million individuals are chronically infected with hepatitis B virus (HBV) globally, resulting in at least 500,000 deaths annually from cirrhosis and hepatocellular carcinoma (HCC) [1]. Africa is heavily burdened by HBV, with an estimated 75 million chronically infected individuals living in the region [1]. Many populations on the continent are also impacted by a high prevalence of co-endemic HIV [2], and around 2.6 million individuals in Africa are living with HBV/HIV coinfection. The KwaZulu-Natal (KZN) province in South Africa has a high HIV prevalence; a 2019 national antenatal survey reported the region to have the highest prevalence in South Africa at 40.9% [3]. HBV prevalence in the region was estimated to be 4.0% in a 2019 household survey, although prevalence was over two-fold higher among HIV positive individuals [4].

HIV co-infection can negatively impact HBV outcomes through potential associations with poorer prognosis, and an increased risk of HBV mother-to-child-transmission (MTCT). However, coinfected individuals can also potentially benefit from shared treatment with nucleos/tide analogue (NA) reverse transcriptase (RT) inhibitors including Lamivudine (3TC), Tenofovir disoproxil fumarate (TDF) and Emtricitabine (FTC) [5,6]. HIV co-infected women are considered to have a 2.5-fold greater risk of HBV MTCT [7,8], although they typically remain on antiretroviral therapy (ART) throughout their pregnancies and this may suppress HBV infection making this risk association less certain [6].

HBV infection outcomes vary considerably with age, with infection in infancy resulting in chronic infection in approximately 95% of cases, and the converse in occurring adults[9]. HBV MTCT transmission, and infections early in infancy are therefore associated with high rates of chronic infection resulting in a long-term burden of liver disease [10,11]. Preventing HBV MTCT is therefore an important goal, and is a key factor in achieving the Global Health Sector Strategy on Viral Hepatitis targets, aiming for a 95% reduction in HBV incidence in children by 2030 [12].

Several factors, including high maternal HBV viral loads (VL) and positive HBV e-antigen (HBeAg) status increase the risk of MTCT, but ART during pregnancy reduces the risk [2]. WHO guidelines advocate that NA treatment should be started between weeks 24-28 of gestation when HBV VL >5.3 log_10_ IU/ml [13]. Additional interventions are also recommended, including HBV immunoglobulin (HBIG) in selected infants, alongside universal birth dose vaccine for neonates (given within the first 24hrs of life) [14]. Infant HBV vaccination also prevents many cases of transmission in infancy and childhood, but must be given as a birth dose (BD) for prevention of MTCT (PMTCT) intervention, as the first dose of the multivalent schedule is not given until 6-10 weeks of age, leaving infants at risk of infection in this window [15,16]. Universal infant vaccination was adopted in South Africa in 1995 [17], with WHO estimates from 2021 suggesting that approximately 85% of infants are vaccinated within the first year of life [18].

Provision of HBV MTCT inventions is patchy across Africa, with just 6% of newborns thought to be reached with timely birth-dose vaccinations in 2019 [19], and issues with cost, supply, safety, and cold-chain limiting access to HBIG [20]. Birth dose HBV vaccination has been endorsed in South Africa, but the intervention has not been rolled out, and monovalent HBV vaccine dose within the first 24h of life is not routinely accessible in practice. NA agents used for HBV antenatal prophylaxis are in wide use in many regions of Africa for HIV, with evidence that this strategy is cost effective [21,22]. However, HIV ART is often provided as fixed-dose combinations, and access to monotherapy for HBV can be challenging, while antenatal screening for HBV to identify at-risk pregnancies that would benefit from treatment is scarce [23]. In South Africa, antenatal HBV screening is recommended in by national guidelines (26), but in practice is rarely undertaken.

MTCT of HIV in South Africa has been drastically reduced, leaving those mothers who do transmit HIV to their child being those with the least interaction with healthcare or poorest acceptance of treatment. To investigate the impact of HIV and its treatment on HBV MTCT in KZN, we screened serum samples from a cohort of mothers known to have transmitted HIV to their infants at birth for HBV biomarkers, to investigate prevalence and characteristics of maternal HBV infection.

## Methods

### Cohort and management of HIV and HBV infections

We studied an established cohort of in utero HIV-transmitting mother-child pairs from the Ucwaningo Lwabantwana (‘learning from children’) study, established in KZN in 2015, established to study the impact of very early combination ART (cART) initiation in infants [24]. Local HIV management guidelines were adhered to [25]. Mothers were initiated on lifelong ART at HIV diagnosis, which for most (75%) of the mothers occurred prior or during pregnancy, but virological failure from poor adherence to ART was typical [24]. During the study period, first line HIV treatment evolved from a fixed-dose combination pill containing FTC, TDF and efaravirenz (EFV), to a fixed-dose combination pill containing 3TC, TDF and dolutegravir (DTG). These treatment regimens were also the local recommendation for the treatment of HBV in HIV coinfection [26]. Maternal study IDs were anonymised for publication.

Infants in the study were treated within the first 48 hrs of life for HIV PMTCT with zidovudine (AZT) or/and nevirapine (NVP) from birth, and switched to combination (c)ART within 21 days of life to suppress HIV infection. 3TC, which is active against HBV, was included in all infant cART regimens. Clinical data and blood samples from the mother-child pairs were collected at enrolment (within 21 days of delivery), monthly for 6 months, then 6-monthly thereafter. Infants in this cohort should therefore have received HBV immunisation as part of a multivalent vaccine schedule starting at age 6-10 weeks, but are unlikely to have received a birth dose vaccination. HBIG is not routinely available in South Africa.

### HBV serology testing

Plasma samples were stored in 0.5ml aliquots at -80°C. Prevalence of maternal HBV infection was determined by the presence of HBsAg, and previous infection by the presence of anti-HBc without HBsAg. HBV vaccination was assumed if anti-HBs was present in the absence of any other HBV biomarker (Table 1). Mothers testing positive for HBsAg were further tested for HBV e-antigen (HBeAg) and HBV DNA quantification in order to determine their risk of MTCT (see Suppl Figure 1 for an outline of the testing approach). Limited volume was available for the infant samples, so infant samples were only screened for HBsAg if the mother was HBsAg-positive. The infants’ samples were screened for HBsAg at 1 and 12 months of age. If at either time point, they were HBsAg positive, HBV DNA was also tested. Anti-HBs was assessed in the infants at 12 months of age, to look for evidence of vaccination.

**Table 1.**
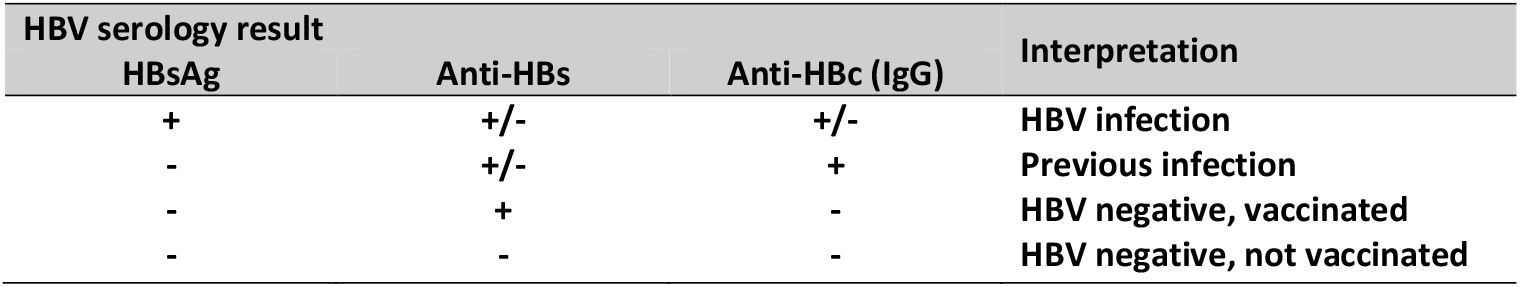
Combinations of HBV serology result and the clinical interpretation. Individuals in the study were also tested for anti-HBc IgM, which is typically considered a marker for acute infection although it is also observed in reactivated infections.

| HBV serology result |  |  | Interpretation |
| --- | --- | --- | --- |
| HBsAg | Anti-HBs | Anti-HBc (IgG) |  |
| + | +/- | +/- | HBV infection |
| - | +/- | + | Previous infection |
| - | + | - | HBV negative, vaccinated |
| - | - | - | HBV negative, not vaccinated |

HBV serology and viral load testing was carried out by Neuberg Global Laboratories in Durban, South Africa. The lower limit of detection for the HBV DNA assay was 1.9 log_10_ IU/ml with an upper limit of quantification of 8.2 log_10_ IU/ml. Samples below the limit of detection were considered HBV DNA negative and those above were assigned a viral load of 8.2 log_10_ IU/ml for statistical analysis. Higher risk of MTCT was assumed in mothers who were HBeAg-positive or with HBV DNA >5.3log_10_ IU/ml (2,3). Further data on maternal age, ART, HIV VL and CD4 count were available for analysis, and were collected as part of the HIV early intervention study. HIV DNA assays had a lower limit of detection of 1.3 log_10_ IU/ml. Statistical analysis was performed using STATA v16.1 and Graphpad Prism v9.4, using Kruskal–Wallis tests to assess differences within the cohort.

### Ethics and Consent

The study was approved by the KwaZulu-Natal Bioethics Research Ethics Committee and the Oxfordshire Research Ethics committees (BF450/14). Written informed consent for the infant and mother’s participation in the study was obtained from the mother or infant’s legal guardian.

## Results

### High maternal HBsAg prevalence is present in this population

A total of 175 HIV-positive mothers were included from the Ucwaningo Lwabantwana study, sampled between July 2015 and April 2021, of whom 8.6% (n=15) were HBsAg positive (Figure 1A). ART was prescribed to 144/175 (82.3%) during pregnancy with 71/175 (40.2%) on ART for at least half their pregnancy. Among HBsAg-positive women, 7/15 (46.7%) were on HBV active ART for <30% of their pregnancies, with three women not listed as being on ART during pregnancy (Table 2). A further 3/15 (20%) women were on treatment for approximately 50% of their pregnancy, and 5/15 (33.3%) were recorded as being on ART for the entire duration. However, as evidenced by vertical HIV transmission in all cases, ART was evidently absent or inconsistent due to numerous social vulnerabilities in these women, as previously described [27].

**Figure 1.**
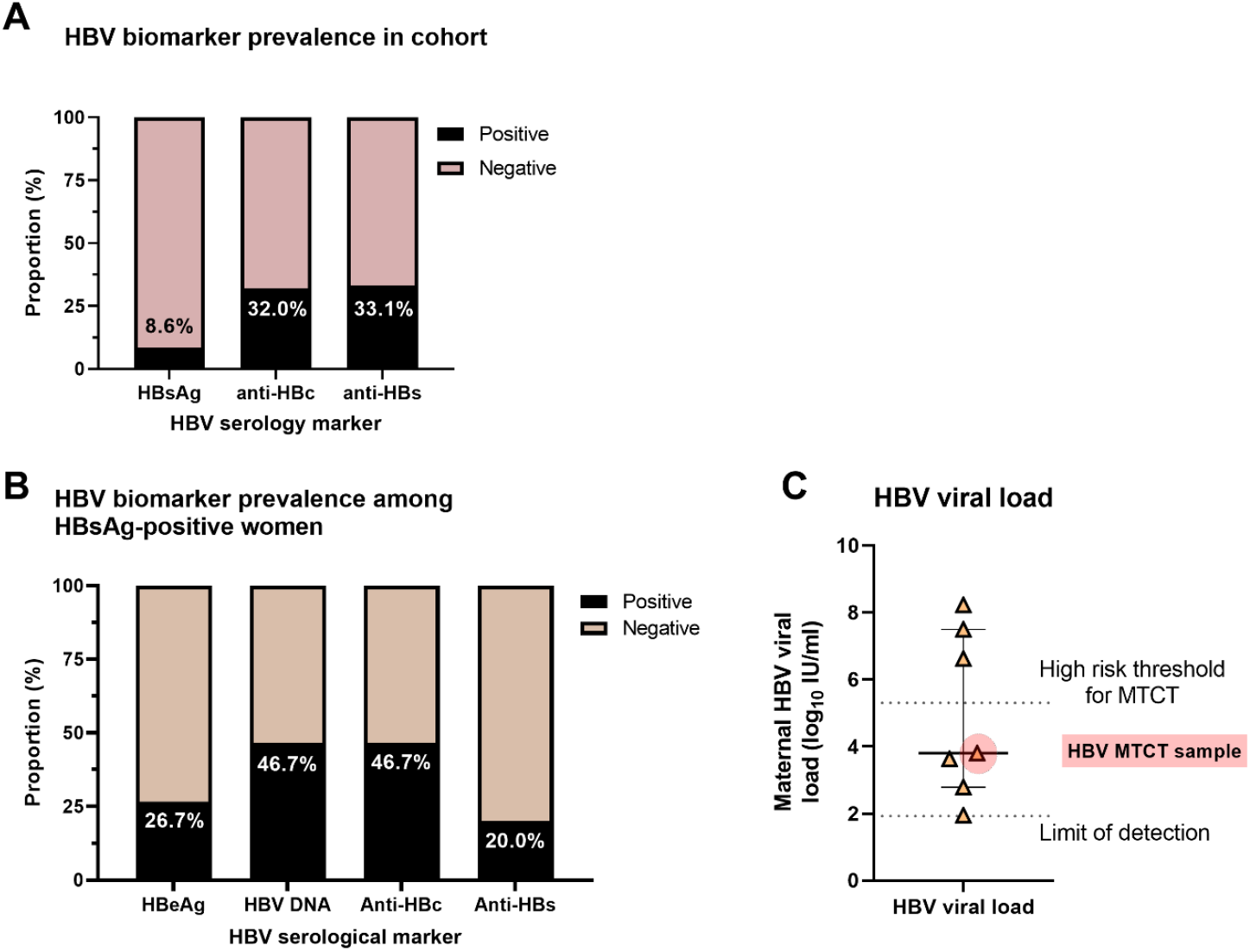
HBV serological profiles of mothers in a cohort of HIV mother-to-child-transmission pairs in KZN, South Africa (A) all women in the cohort (n=175), (B) all women testing HBsAg-positive (n=15) and (C) viral loads of the HBV DNA positive women (n=7). All women in the cohort were tested for HBV surface antigen (HBsAg), HBV anti-core IgG (anti-HBc) and HBV anti-S (anti-HBs). In (B) additional testing of HBsAg-positive women is presented, which included HBV e-antigen (HBeAg) and HBV DNA. Seven of the HBsAg-positive women had HBV viral loads above the limit of detection (1.92log_10_ IU/ml), and these are plotted in (C) with the median and interquartile range shown. The viral load of the maternal sample from the HBV MTCT pair is highlighted in red. The high-risk threshold for mother to child transmission (MTCT) of HBV is also indicated (5.3log_10_ IU/ml).

**Table 2.** Maternal HBV and HIV virology and serology results among HBsAg-positive (n=15) and anti-HBc IgM positive (n=1) women and their infants. Data on the ART regimen prescribed and the proportion of the pregnancy on ART is included, with HBsAg screening results for the infants at 1 and 12 months. HBV mother to child transmission (MTCT) risk was based on maternal HBV viral load, with the WHO threshold of >5.3log10 IU/ml used to identify high risk pregnancies. One mother (ID HBKN-109) was HBsAg negative, and anti-HBc IgM positive suggesting either recent or reactivating infection. As interpretation was unclear, samples from the paired infant were screened. One mother-child pair had no infant samples available for testing, another sample had insufficient volume at 12 months, and another was lost to follow up, meaning no 12-month sample was available to test (not available; NA)

| Maternal study ID | Maternal HBV serology and virology |  |  |  |  | Maternal HIV viral load, CD4 and CD8 counts, and treatment at delivery |  |  |  |  | Infant HBV serology |  |  | Infant HIV treatment |  |  |
| --- | --- | --- | --- | --- | --- | --- | --- | --- | --- | --- | --- | --- | --- | --- | --- | --- |
| | Anti-HBs | Anti-HBc | HBeAg | Maternal HBV viral load ($\log_{10}$ IU/ml) | HBV MTCT risk | Proportion pregnancy on ART (%) | HBV active NA in maternal Treatment* | Maternal HIV VL ($\log_{10}$ copies/ml) | CD4 (cells/ml) | CD8 (cells/ml) | Infant HBsAg 1 month | Infant HBsAg 12 months | Infant anti-HBs 12 months | Infant PMTCT at birth** | Infant cART** | Age at cART initiation (days)† |
| HBKN-109 | + | + [IgM+] | n/a | n/a | Unclear | 69.5 | TDF | 3.3 | 653 | 1554 | - | - | + | AZT | 3TC/AZT/NVP | 8-14 |
| HBKN-26 | - | + | - | 1.94 | Low | 100.0 | TDF | <1.30 | 479 | 1089 | - | - | + | NVP/AZT | AZT/3TC/NVP | ≤7 |
| HBKN-61 | - | - | - | <1.92 | Low | 45.9 | TDF | <1.30 | 552 | 1358 | - | - | - | AZT/NVP | AZT/3TC/NVP | 8-14 |
| HBKN-62 | + | + | - | <1.92 | Low | 4.9 | TDF | 3.53 | 481 | 1717 | - | - | + | NVP | AZT/3TC/NVP | ≤7 |
| HBKN-81 | - | - | - | <1.92 | Low | 6.3 | TDF | 2.93 | 1183 | 946 | - | - | + | NVP/AZT | 3TC/AZT/NVP | 8-14 |
| HBKN-99 | - | + | - | 3.8 | Low | 100.0 | 3TC | 5.64 | 331 | 727 | - | + | + | NVP/AZT | 3TC/AZT/NVP | 15-21 |
| HBKN-100 | + | + | - | <1.92 | Low | 0.0 | TDF | 4.59 | 606 | 1349 | - | - | + | NVP/AZT | 3TC/AZT/NVP | ≤7 |
| HBKN-103 | + | + | - | <1.92 | Low | 0.0 | TDF | 3.45 | 437 | 2213 | - | - | - | NVP/AZT | 3TC/AZT/NVP | ≤7 |
| HBKN-114 | - | - | - | <1.92 | Low | 100.0 | TDF | 5.11 | 770 | 806 | NA | NA | NA | AZT | 3TC/AZT/NVP | ≤7 |
| HBKN-117 | - | + | - | <1.92 | Low | 2.9 | TDF | 4.84 | 726 | 1154 | - | - | + | AZT/NVP | 3TC/ABC/KAL | ≥22 |
| HBKN-149 | - | - | - | 3.63 | Low | 12.4 | TDF | 4.95 | 333 | 1273 | - | - | + | NVP | 3TC/NVP/AZT | 8-14 |
| HBKN-160 | - | - | - | <1.92 | Low | 0.0 | TDF | <1.30 | 389 | 967 | - | - | - | AZT/NVP | 3TC/AZT/NVP | 8-14 |
| HBKN-72 | - | - | + | 2.78 | Low | 57.5 | 3TC | 4.08 | 50 | 442 | - | NA | NA | NVP/AZT | 3TC/AZT/NVP | ≤7 |
| HBKN-05 | - | - | + | >8.22 | High | 100.0 | TDF | 5.40 | 252 | 777 | - | NA | NA | NVP/AZT | 3TC/NVP/AZT | ≤7 |
| HBKN-118 | - | + | + | 6.62 | High | 53.8 | TDF | <1.30 | 292 | 722 | - | - | - | AZT/NVP | 3TC/AZT/NVP | 15-21 |
| HBKN-159 | - | - | + | 7.49 | High | 100.0 | TDF | 4.60 | 109 | 375 | - | - | - | AZT/NVP | 3TC/AZT/NVP | ≤7 |

Proportion of pregnancy on ART (%)
\*Maternal treatments included TDF (tenofovir) or 3TC (lamivudine). \*\*Infant HIV PMTCT included AZT (zidovudine) or/and NVP (nevirapine). Combination (c)ART also included 3TC, ritonavir boosted lopinavir (LPV/r; referred to as Kaletra, KAL) and abacavir (ABC). NA – sample not available. † The study aimed to prescribe infant cART within 21 days of life

### High risk of HBV MTCT is present despite prescription of HBV-active ART

Among HBsAg-positive women, 4/15 (26.7%) were HBeAg-positive and 7/15 (46.7%) were HBV DNA positive (Table 2, Figure 1B). All HBeAg-positive women had detectable HBV DNA, with 3/4 of these women having VL classified as high risk for MTCT (>5.3log_10_ IU/ml) (Figure 1C). Median HBV VL among those with detectable viraemia was 3.8log_10_ IU/ml. HBsAg-positive women were younger (mean 21.7yrs) than HBsAg-negative women (mean 25.5yrs) (p=0.009, Table 3). No difference in age was associated with the presence of HBV DNA (Table 2). Age was also associated with HBV exposure in the cohort, with anti-HBc negative women being younger (median 22yrs) than anti-HBc positive women (median 26yrs) (p<0.001, Suppl Fig 2).

**Table 3.** Age and HIV associated factors and association with HBsAg and presence of HBV DNA. The age of the mothers in the cohort, proportion of their pregnancy on antiretroviral therapy (ART), HIV viral loads (VL) and CD4 counts are given for HBsAg positive and negative women, and HBV DNA positive and negative women.

| Among KZN Ucwningo Lwabantwana mothers (n=175) |  |  |  |  |  |
| --- | --- | --- | --- | --- | --- |
|  | HBsAg positive (n=15) |  | HBsAg negative (n=160) |  |  |
|  | Mean | 95%CI | Mean | 95%CI | p |
| Age (yrs) | 21.7 | 20.3-23.2 | 25.5 | 24.7-26.4 | 0.009 |
| Pregnancy on ART (%) | 45.6 | 23.0-68.1 | 43.4 | 37.4-49.5 | 0.927 |
| HIV VL (log<sub>10</sub> copies/ml) | 4.9 | 4.0-5.1 | 4.9 | 4.6-5.1 | 0.733 |
| CD4 count (cells/mm<sup>3</sup>) | 466 | 321.8-610.2 | 518.8 | 473.6-564.0 | 0.481 |
| Among HBsAg positive mothers only (n=15) |  |  |  |  |  |
|  | HBV DNA positive (n=7) |  | HBV DNA negative (n=8) |  |  |
|  | Mean | 95%CI | Mean | 95%CI | p |
| Age (yrs) | 21.5 | 18.8-24.3 | 22.0 | 19.8-24.2 | 0.745 |
| Pregnancy on ART (%) | 74.8 | 42.8-100.0 | 20.0 | 0.0-49.9 | 0.011 |
| HIV VL (log<sub>10</sub> copies/ml) | 4.0 | 2.2-5.6 | 3.4 | 2.1-4.6 | 0.354 |
| CD4 count (cells/mm<sup>3</sup>) | 263.7 | 129.6-397.9 | 643.0 | 429.3-856.7 | 0.003 |

### Anti-core (anti-HBc) antibody is an unreliable marker of exposure in this HIV-positive population

Approximately a third of mothers in the study were anti-HBc positive (32.0%, 56/175), indicating serological evidence of exposure to HBV. However, among the HBV-positive women, only 7/15 HBsAg-positive samples were IgG anti-HBc positive (46.7%). IgM anti-HBc was positive for an additional mother, who was HBsAg negative but was also anti-HBs and IgG anti-HBc positive, potentially suggesting recent exposure and clearance, or reactivation.

### Serological evidence of vaccine-mediated HBV immunity is present in a minority of women

Anti-HBs is generally considered to be a marker of protective immunity, and can be induced either by vaccination or by natural infection. Anti-HBs was detected in 33.1% mothers (58/175) (Figure 1). Anti-HBs was rare among the HBsAg-positive women, with just 20% of these women (n=3) testing anti-HBs positive. These three women were all anti-HBc positive, HBeAg negative and HBV DNA was below the limit of detection.

To estimate the prevalence of HBV vaccination within the cohort, we identified women who were anti-HBs positive, but negative for both anti-HBc and HBsAg (Figure 2). Among 58 women who were anti-HBs positive, 44 (75.9%) had evidence of another HBV biomarker and only 14/58 women had an anti-HBs only profile, reflecting a low prevalence of vaccine-mediated immunity, at just 8.0% in the cohort overall. The presence of anti-HBs was not associated with HIV VL or CD4 count (Suppl Figure 3).

**Figure 2.**
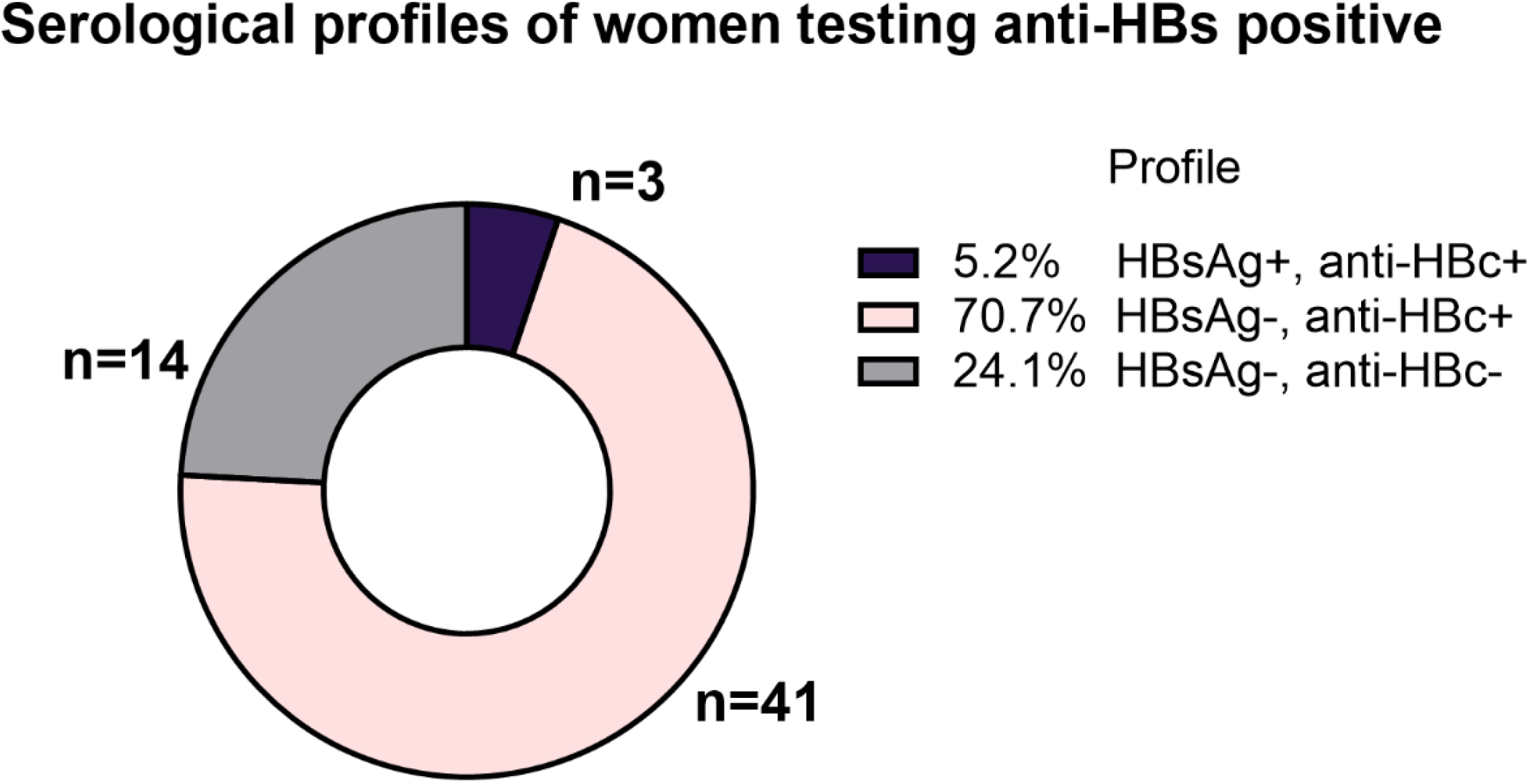
Serological profiles of women testing HBV anti-S (anti-HBs) positive (n=58). Anti-HBs is generally considered to be a marker of protective immunity for HBV, and can be either vaccine induced or develops subsequent to HBV infection. HBV vaccination was likely in HBsAg- / anti-HBc-women, with the presence of any other HBV biomarker (HBsAg and/or anti-HBc) suggesting immunity is the result of previous infection.

### The prescription of maternal ART did not correlate with HBV or HIV aviraemia

HIV viraemia was common among all the mothers (median 4.9 log_10_ HIV RNA copies/mL), as expected for HIV transmitting mothers. Comparing HBsAg-positive and -negative women, there were no differences in HIV VL, CD4 or CD8 count (Suppl fig 4). Among HBsAg-positive women recorded as being on ART for less than a trimester (n=7), the majority (6/7) had undetectable HBV DNA, although 4/7 had CD4 counts <500 cells/mm^3^ and 6/7 women had detectable HIV viraemia (Table 2). HIV viraemia was undetectable in 4/15 HBsAg-positive mothers, and three of these women also had HBV VL near or below the limit of detection. CD4 counts were lower in HBV DNA positive women than in their virologically suppressed counterparts (263.7 cells/mm^3^ vs 643.0 cells/mm^3^) (Table 3). One mother (ID HBKN-118) had a HBV VL 6.6 log_10_ IU/ml, and was aviraemic for HIV, indicating a similar clinical phenotype to previously reported cases of HBV drug resistance in South Africa where suppression of HIV is a proxy of good adherence to therapy [28]. Women with detectable HBV DNA had spent a greater proportion of their pregnancy on ART (74.8% vs 20.0%, p=0.011), although as noted there were issues with adherence.

### HBV detected in one infant at age 12 months in a mother with detectable HBV and HIV viraemia at birth

The infants of HBV-positive mothers were screened for HBsAg at 1- and 12-months of age where samples were available (Table 2). All screened infants were HBV-negative at 1 month (n=14; no sample for one infant), and 1/13 (7.7%) infant (maternal ID HBKN-99) was HBV-positive at 12 months with HBV VL.3.1 log_10_ IU/ml. Samples were unavailable for three infants at 12 months of age. At birth, the mother of the HBsAg-positive infant was HBeAg-negative with HBV VL 3.8 log_10_ IU/ml, and was recorded as taking TDF-based therapy throughout pregnancy but had a detectable HIV VL of 5.6 log_10_ copies/ml. Of note, this infant was not commenced on 3TC containing cART until 21 days of life, compared with the overall median of 6.5 days (IQR 2.3 – 12.8) among other infants. Despite the ongoing prescription of cART, at 12-months the infant had an HIV VL of > 8 log_10_ HIV RNA copies/mL and did not achieve HIV plasma viral suppression until 18 months of age, suggesting poor adherence. HBV infection was not detected in any of the infants of the mothers deemed high transmission risk. The infant from the IgM anti-HBc positive mother was also screened at 1 month and 12 months, and both of these infant samples tested negative (Table 2).

### Infants with vertical HIV infection have a low rate of vaccine-mediated immunity to HBV infection

All available 12-month infant samples that were screened for HBsAg were also screened for anti-HBs (n=13; including the infant of the IgM-positive mother), to look for evidence of HBV vaccination (Table 2). An anti-HBs only serological profile was observed in 7/13 (53.8%) infants. The HBsAg-positive infant also tested anti-HBs-positive (1/13), and the remaining samples were anti-HBs negative (5/13). This suggests that only 7/13 infants in this cohort received infant vaccination, although we cannot exclude the possibility either that partial vaccination series were delivered, or that despite vaccination, anti-HBs titres were not mounted due to co-existing HIV infection.

## Discussion

The HBV prevalence of 8.6% in this vulnerable cohort of HIV-transmitting mothers, is classified as ‘high’ based on WHO thresholds. Among the coinfected group, 20% are considered high-risk for HBV MTCT based on virologic parameters, reaching this threshold despite having availability of antiviral agents active against HBV. Despite these risks, all infants from HBV infected mothers in the cohort tested HBV negative at birth.

A single HBV infection was identified in an infant at 12 months of age, born to a mother who was not classified as ‘high risk’ based on WHO guidelines [13]. However, all HIV positive women should be prescribed ART regardless of HBV VL [29,30] and poorly controlled HIV infection is itself a risk factor for HBV transmission in this case. It is noteworthy that the majority of data informing these guidelines stem from Asia [31], and that studies in Africa have documented HBV MTCT at lower HBV DNA VL and in HBeAg-negative women [32,33].

The infants were part of an early intervention study for HIV, and were all treated with cART within 21 days of birth [24]. Whilst the ART regimens used at birth did not typically contain compounds active against HBV, cART containing 3TC was initiated at a median of 6.5 days. It is possible this early invention for HIV may have also minimised HBV MTCT events in this cohort. Approximately half the infants tested had serology consistent with vaccination which would have occurred from 6 weeks as per the local guidelines in line with the WHO Expanded Program on Immunization (EPI). Serology suggested a lack of vaccine-mediated immunity in 5/13 infants tested, suggesting a high proportion of infants remain at risk of horizontal transmission later in childhood. The case of HBV MTCT identified, and the low prevalence of protective anti-HBs antibody, highlight that many infants remain at risk of HBV infection in early life, and our data underscore the importance of combining antenatal HBV screening with maternal prophylaxis, and infant vaccination starting with a timely birth dose and follow-up doses during infancy.

Anti-HBc has been used previously in epidemiological models of HBV transmission; in HBsAg-negative individuals it represents exposure to (and recovery from) HBV infection [34,35]. IgM anti-HBc appears early in infection, before isotype-switching to IgG when chronic infection establishes or the infection is cleared [36]. IgG anti-HBc is assumed to persist for life, making it useful for population-based screening [37]. Over half the women who were HBsAg positive in this cohort were also anti-HBc negative, with recent infection unlikely. This suggests that HIV infection is compromising the generation of anti-HBC antibodies in these women. This serologic profile has also been described in other co-infected cohorts [38,39], associated with HIV (especially with low CD4 counts) and in patients undergoing solid organ transplants, suggesting an association with immune compromise [40]. In our study, there was no difference in the CD4 or CD8 counts between women who were anti-HBc negative/HBsAg-positive and anti-HBc positive (Suppl Figure 5). This HBsAg-positive/anti-HBc negative profile suggests that in populations where HIV is co-endemic, screening based on anti-HBc may underestimate the proportion of the population exposed, and approaches to mitigate for this should be considered.

Evidence of maternal HBV vaccination was low in this study, with just 8.0% having a serological profile consistent with HBV vaccination. However, it is possible that a higher proportion received childhood vaccination but that vaccine-mediated antibody titres had waned over time, which is recognised particularly in the context of HIV infection [34]. Increased vaccination boosters, in conjunction with screening for high-risk adult populations such as these, could be considered to reduce the risk of MTCT. Effective HBV vaccination among HIV-positive individuals remains challenging however, with logistical difficulties in identifying those eligible and delivering a three-dose vaccine schedule in adulthood, and vaccine efficacy dropping to 35-70% in this population [41]. There remains a need for more immunogenic vaccines and further work to optimise the boosting schedule for the vaccine within immunocompromised populations [42].

HBsAg-positive women were on average four-years younger than other women in the cohort, suggesting younger women in the cohort were particularly vulnerable to HBV MTCT. Previous research has indicated adolescent and younger mothers are generally less engaged with the HIV continuum of care for PMTCT than older women, placing their infants at greater risk of HIV transmission [43,44]. These increased risks may also apply to HBV MTCT.

## Limitations

The number of HBV-positive women identified in the study was small, making it difficult to generate robust conclusions of the impact of early treatment intervention on the prevention of HBV MTCT. A single mother-child infected pair was identified. HBV sequencing to confirm the mother and child’s infection were closely related would have been helpful, but both had a relatively low HBV DNA (∼3.0 log_10_ IU/ml), which makes sequencing challenging [45]. Sequencing would also have been informative to better understand the clinical phenotype in the mother who had undetectable HIV but a high HBV viral load. Whilst the presentation can suggest an ART resistant HBV infection, the mother was only on ART for little over half her pregnancy and HBV VL typically take longer to decline on treatment than HIV [46], so it is possible the therapy had not had sufficient time to suppress HBV DNA.

Occult HBV, where individuals are HBsAg-negative but HBV DNA positive [47], has been reported to be more common among HIV-positive individuals [48]. The testing algorithm used in our study did not screen for occult HBV, and it is therefore possible that a number of HBV cases were overlooked. Further work on the prevalence of occult HBV, and the potential risk of HBV MTCT in these cases would be informative. Infant samples were only screened for HBsAg and anti-HBs, with the presence of anti-HBs taken as evidence of HBV vaccination. Anti-HBc would have been informative to rule out exposure to HBV also, but as no infants were HBV-positive at birth, a HBsAg-negative, anti-HBs profile was considered sufficient.

## Conclusions

Findings from this study indicate a high level of HBV prevalence among mothers living with HIV in KZN, putting their pregnancies at risk of both HIV and HBV MTCT. It is important to avoid complacency in assuming that prescription of ART active against both HIV and HBV prevents against transmission events. However, maternal ART and the early ART treatment of infants likely contributed to reducing the chances of infant HBV infections in high-risk mothers. A role for both the increased use of maternal prophylaxis and potential neonatal post-exposure prophylaxis in high-risk infants, in addition to birth dose and continued infant HBV vaccination, should be considered. Interdisciplinary interventions are required to support disadvantaged women who are at high risk of transmitting both HBV and HIV to their infants, to enhance screening, adherence to ART regimens throughout pregnancy, and administration of timely birth-dose HBV vaccination. These interventions are crucial to support progress towards international elimination goals for HBV.

## Data Availability

HBV serology data for all 175 mothers and infants tested as a part of the study are available on Figshare: https://doi.org/10.6084/m9.figshare.22259224
Findings from this study were presented as a poster presentation at the EASL International Liver Congress, London, 2022 and the poster can be viewed via this link: https://doi.org/10.6084/m9.figshare.22118381.v1

https://doi.org/10.6084/m9.figshare.22259224

https://doi.org/10.6084/m9.figshare.22118381.v1

## Acknowledgments

We would like to acknowledge all the women and their infants who have participated in the Ucwaningo Lwabantwana (‘learning from children’) study over the years, and the many healthcare workers who have supported the families throughout.

## Funding

This study was funded by an Oxford University John Fell Fund award (ref. 0008410) held by Anna McNaughton. Supplementary testing was funded by the Wellcome Trust. Philippa Matthews is funded by Wellcome (grant ref 110110/Z/15/Z), the Francis Crick Institute and UCLH NIHR Biomedical Research Centre (BRC).

All authors contributing to the study were given the opportunity to comment on the manuscript, and approved the final, submitted version of the manuscript.

## Data availability

HBV serology data for all 175 mothers and infants tested as a part of the study are available on Figshare: https://doi.org/10.6084/m9.figshare.22259224

Findings from this study were presented as a poster presentation at the EASL International Liver Congress, London, 2022 and the poster can be viewed via this link: https://doi.org/10.6084/m9.figshare.22118381.v1

## Additional Figures in supplementary data file

Supplementary figure 1; Testing approach used to screen mothers and infants in the HBV study, indicating when further testing was required.

Supplementary Figure 2; Ages of mothers in the cohort and anti-HBc status.

Supplementary Figure 3; (A) CD4 counts and (B) HIV viral loads stratified by anti-HBs status.

Supplementary Figure 4; (A) HIV viral load, (B) CD4 count and (C) CD8 count of mothers stratified by HBsAg status.

Supplementary Figure 5; (A) CD4 counts and (B) CD8 counts stratified by anti-HBc status.

## References

1. Sheena BS, Hiebert L, Han H, et al. Global, regional, and national burden of hepatitis B, 1990– 2019: a systematic analysis for the Global Burden of Disease Study 2019. The Lancet Gastroenterology & Hepatology. 2022; 1253(22):1990–2019.

2. Matthews PC, Geretti AM, Goulder PJR, Klenerman P. Epidemiology and impact of HIV coinfection with Hepatitis B and Hepatitis C viruses in Sub-Saharan Africa. Journal of Clinical Virology. Elsevier B.V.; 2014; 61:20–33.

3. Woldesenbet SA, Lombard C, Manda S, et al. The 2019 National Antenatal HIV Sentinel Survey, South Africa. National Department of Health. 2021;.

4. Samsundera N, Ngcapua S, Lewisa L, et al. Seroprevalence of hepatitis B virus: Findings from a population-based household survey in KwaZulu-Natal, South Africa. International Journal of Infectious Diseases. 2019; 85:150–157.

5. Villa G, Iwuji C. HIV and chronic hepatitis B virus co-infection in sub-Saharan Africa: a deadly synergy. Public Health Action. 2020; 10(2):31–49.

6. Maponga TG, McNaughton AL, Van Schalkwyk M, et al. Treatment advantage in HBV/HIV coinfection compared to HBV monoinfection in a South African cohort. Journal of Infection [Internet]. Elsevier Ltd; 2020; (xxxx). Available from: http://dx.doi.org/10.1016/j.jinf.2020.04.037

7. Andersson MI, Maponga TG, Ijaz S, Theron G, Preiser W, Tedder RS. High HBV viral loads in HIV-infected pregnant women at a tertiary hospital, South Africa. J Acquir Immune Defic Syndr. Ovid Technologies (Wolters Kluwer Health); 2012; 60(4):e111–2.

8. Hoffmann CJ, Thio CL. Clinical implications of HIV and hepatitis B co-infection in Asia and Africa. Lancet Infect Dis. Elsevier BV; 2007; 7(6):402–409.

9. World Health Organization. Hepatitis B factsheet. 2022; :https://www.who.int/news-room/fact-sheets/detail/h.

10. Andersson MI, Rajbhandari R, Kew MC, et al. Mother-to-child transmission of hepatitis B virus in sub-Saharan Africa: Time to act. The Lancet Global Health. Andersson et al. Open Access article distributed under the terms of CC BY; 2015; 3(7):e358–e359.

11. Wilson P, Parr JB, Jhaveri R, Meshnick SR. Call to Action: Prevention of Mother-to-Child Transmission of Hepatitis B in Africa. Journal of Infectious Diseases. 2018; 217(8):1180–1183.

12. World Health Organization. Global health sector strategy on viral hepatitis 2016–2021. 2016.

13. WHO. Prevention of Mother-to-Child Transmission of Hepatitis B Virus: Guidelines on Antiviral Prophylaxis in Pregnancy. 2020.

14. Nayagam S, Shimakawa Y, Lemoine M. Mother-to-child transmission of hepatitis B: What more needs to be done to eliminate it around the world? Journal of Viral Hepatitis. 2020; 27(4):342–349.

15. Bodo B, Malande OO. Delayed introduction of the birth dose of Hepatitis B vaccine in EPI programs in East Africa: a missed opportunity for combating vertical transmission of Hepatitis B. The Pan African medical journal. 2017; 27(Supp 3):19.

16. Johannessen A, Mekasha B, Desalegn H, Aberra H, Stene-Johansen K, Berhe N. Mother-to-child transmission of hepatitis B virus in Ethiopia. Vaccines. 2021; 9(5):1–10.

17. Burnett RJ, Kramvis A, Dochez C, Meheus A. An update after 16 years of hepatitis B vaccination in South Africa. Vaccine. Netherlands; 2012; 30 Suppl 3:C45–51.

18. Stone J, Artenie A, Hickman M, et al. The contribution of unstable housing to HIV and hepatitis C virus transmission among people who inject drugs globally, regionally, and at country level: a modelling study. The Lancet Public Health. The Author(s). Published by Elsevier Ltd. This is an Open Access article under the CC BY 4.0 license; 2022; 7(2):e136–e145.

19. Villiers M de, Nayagam S, Hallett TB. The impact of the timely birth-dose vaccine on the global elimination of hepatitis B. Nature Communications. Springer US; 2021; (12):6223.

20. Dionne-Odom J, Njei B, Tita ATN. Elimination of Vertical Transmission of Hepatitis B in Africa: A Review of Available Tools and New Opportunities. Clinical Therapeutics. 2018; 40(8):1255–1267.

21. Tamandjou Tchuem CR, Andersson MI, Wiysonge CS, Mufenda J, Preiser W, Cleary S. Prevention of hepatitis B mother-to-child transmission in Namibia: A cost-effectiveness analysis. Vaccine. The Authors; 2021; 39(23):3141–3151.

22. Mokaya J, Burn EAO, Tamandjou CR, et al. Modelling cost-effectiveness of tenofovir for prevention of mother to child transmission of hepatitis B virus (HBV) infection in South Africa. BMC Public Health. BMC Public Health; 2019; 19(1):1–9.

23. Spearman CW, Afihene M, Ally R, et al. Hepatitis B in sub-Saharan Africa: strategies to achieve the 2030 elimination targets. The Lancet Gastroenterology & Hepatology. Netherlands; 2017; 2(12):900–909.

24. Millar JR, Bengu N, Vieira VA, et al. Early Initiation of Antiretroviral Therapy Following In Utero HIV Infection Is Associated With Low Viral Reservoirs but Other Factors Determine Viral Rebound. Journal of Infectious Diseases. 2021; 224(11):1925–1934.

25. Republic of South Africa National Department of Health. 2019 ART Clinical Guidelines for the Management of HIV in Adults, Pregnancy, Adolescents, Children, Infants and Neonates [Internet]. Available from: https://www.health.gov.za/wp-content/uploads/2020/11/2019-art-guideline.pdf

26. Chotun N, Preiser W, Rensburg CJ van, et al. Point-of-care screening for hepatitis B virus infection in pregnant women at an antenatal clinic: A South African experience. PLoS One. Public Library of Science (PLoS); 2017; 12(7):e0181267.

27. Millar JR, Bengu N, Fillis R, et al. High-frequency failure of combination antiretroviral therapy in paediatric HIV infection is associated with unmet maternal needs causing maternal non-adherence. EClinicalMedicine. Elsevier Ltd; 2020; 22:100344.

28. Mokaya J, Maponga TG, McNaughton AL, et al. Evidence of tenofovir resistance in chronic hepatitis B virus (HBV) infection: An observational case series of South African adults. Journal of Clinical Virology. Elsevier; 2020; 129(June):104548.

29. European Association for Study of the Liver. EASL 2017 Clinical Practice Guidelines on the management of hepatitis B virus infection. Journal of Hepatology. 2017; 67(2):370–398.

30. World Health Organization. Guidelines for the Prevention, Care and Treatment of Persons with Chronic Hepatitis B Infection. Who. 2015; (March):166.

31. Funk AL, Lu Y, Yoshida K, et al. Efficacy and safety of antiviral prophylaxis during pregnancy to prevent mother-to-child transmission of hepatitis B virus: a systematic review and meta-analysis. The Lancet Infectious Diseases. Elsevier; 2021; 21(1):70–84.

32. Shimakawa Y, Veillon P, Birguel J, et al. Residual risk of mother-to-child transmission of hepatitis B virus infection despite timely birth-dose vaccination in Cameroon (ANRS 12303): a single-centre, longitudinal observational study. The Lancet Global Health. 2022; 10(4):e521–e529.

33. Candotti D, Danso K, Allain J-P. Maternofetal transmission of hepatitis B virus genotype E in Ghana, west Africa. Journal of General Virology. 2007; 88(10):2686–2695.

34. McNaughton A, Lourenco J, Hattingh L, et al. HBV vaccination and PMTCT as elimination tools in the presence of HIV: insights from a clinical cohort and dynamic model. BMC Medicine. BMC Medicine; 2019; 17(43):https://doi.org/10.1186/s12916-019-1269-x.

35. Mcnaughton AL, Lourenco J, Bester PA, et al. Hepatitis B virus seroepidemiology data for Africa: Modelling intervention strategies based on a systematic review and meta-analysis. PLoS Medicine. 2020; (vember 2019):1–22.

36. Gish RG, Basit SA, Ryan J, Dawood A, Protzer U. Hepatitis B Core Antibody: Role in Clinical Practice in 2020. Current Hepatology Reports. 2020; 19(3):254–265.

37. Konerman MA, Lok AS. Epidemiology, Diagnosis, and Natural History of Hepatitis B. Seventh Ed. Elsevier Inc.; 2018.

38. Demir M, Phiri S, Heger E, et al. Prevalence of Anti-HBs Without Anti-HBc Among HIV-Infected Adults Initiating Antiretroviral Therapy in Lilongwe, Malawi. Journal of Acquired Immune Deficiency Syndromes. 2018; 78(3):E14–E15.

39. Price H, Dunn D, Zachary T, et al. Hepatitis B serological markers and plasma DNA concentrations. AIDS. England; 2017; 31(8):1109–1117.

40. Avettand-Fenoel V, Thabut D, Katlama C, Poynard T, Thibault V. Immune suppression as the etiology of failure to detect anti-HBc antibodies in patients with chronic hepatitis B virus infection. Journal of Clinical Microbiology. 2006; 44(6):2250–2253.

41. Farooq PD, Sherman KE. Hepatitis B Vaccination and Waning Hepatitis B Immunity in Persons Living with HIV. Current HIV/AIDS Reports. Current HIV/AIDS Reports; 2019; 16(5):395–403.

42. Mohareb AM, Kim AY. Hepatitis B Vaccination in People Living with HIV - If at First You Don’t Succeed, Try Again. JAMA Network Open. 2021; 4(8):2021–2023.

43. Burrage AB, Mushavi A, Shiraishi RW, et al. Mother-To-Child Transmission of HIV in Adolescents and Young Women: Findings From a National Prospective Cohort Survey, Zimbabwe, 2013– 2014. Journal of Adolescent Health. 2020; 66(4):455–463.

44. UNICEF. Addressing the needs of adolescent and young mothers affected by HIV in Eastern and Southern Africa. 2020.

45. McNaughton AL, Roberts HE, Bonsall D, et al. Illumina and Nanopore methods for whole genome sequencing of hepatitis B virus (HBV). Scientific Reports [Internet]. Springer US; 2019; 9(7081). Available from: http://dx.doi.org/10.1101/470633

46. Anderson M, Gaseitsiwe S, Moyo S, et al. Slow CD4+ T-Cell recovery in human immunodeficiency virus/hepatitis B virus-coinfected patients initiating truvada-based combination antiretroviral therapy in Botswana. Open Forum Infectious Diseases. 2016; 3(3):1–8.

47. Raimondo G, Locarnini S, Pollicino T, Levrero M, Zoulim F, Lok AS. Update of the statements on biology and clinical impact of occult hepatitis b virus infection. European Association for the Study of the Liver; 2019.

48. Ryan K, Anderson M, Gyurova I, et al. High Rates of Occult Hepatitis B Virus Infection in HIV-Positive Individuals Initiating Antiretroviral Therapy in Botswana. Open Forum Infectious Diseases [Internet]. 2017; 4(4). Available from: http://dx.doi.org/10.1093/ofid/ofx195

