## Supplementary data file for "Hepatitis B Virus (HBV) prevalence and characteristics in HIV-transmitting mothers and their infants in KwaZulu-Natal, South Africa"

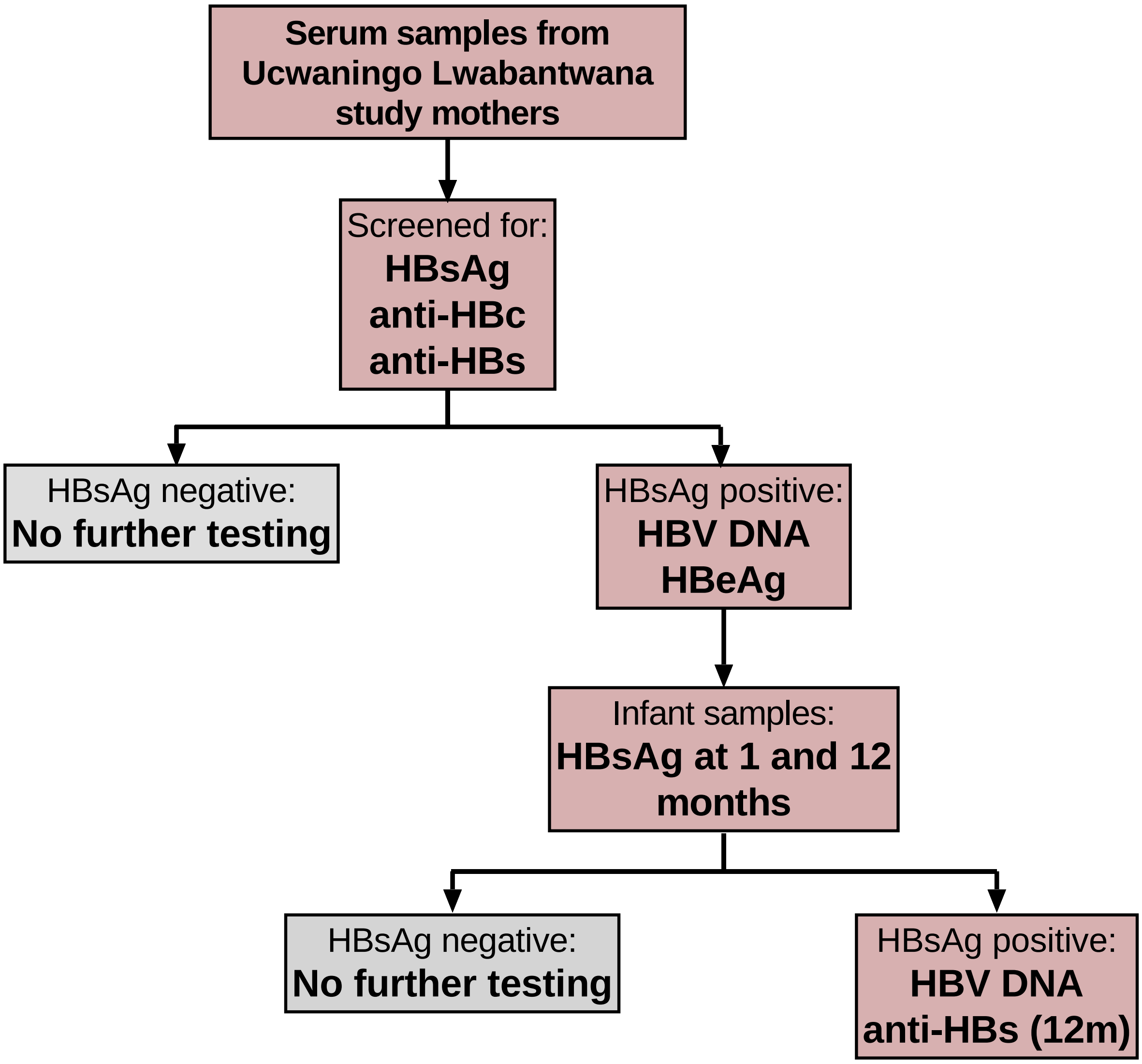


**Supplementary figure 1; Testing approach used to screen mothers and infants in the HBV study, indicating when further testing was required.** Maternal samples were for HBsAg, anti-HBs, anti-HBc IgM and total anti-HBc. Mothers were considered HBV-positive if HBsAg was detected and these samples were further tested for HBV DNA and HBeAg. Only the infants of HBsAg-positive women were tested for HBsAg. Infant samples were only tested for anti-HBs at 12 months of age, as infant vaccination typically occurs after 6 weeks of age.

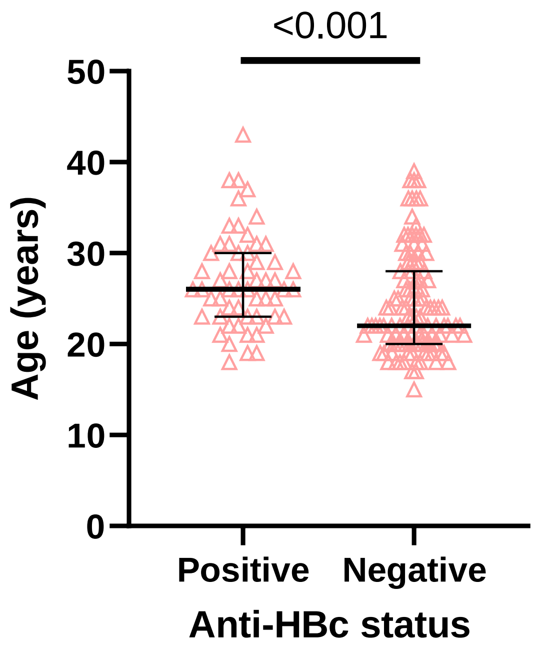


**Supplementary Figure 2; Ages of mothers in the cohort and anti-HBc status.** Median and interquartile ranges are indicated, with p values.

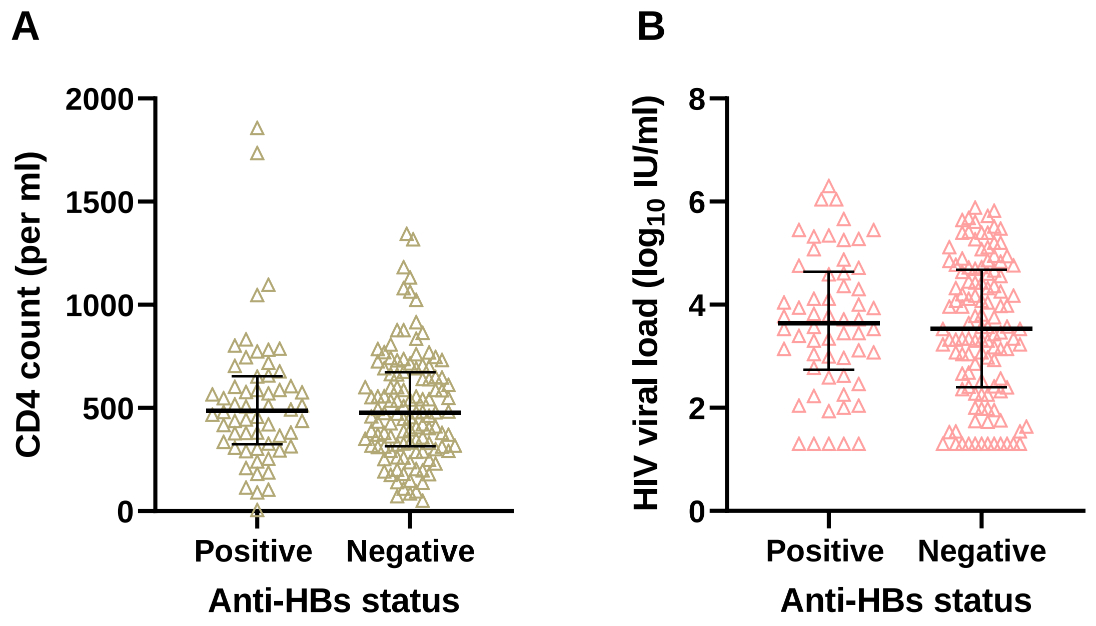


**Supplementary Figure 3; (A) CD4 counts and (B) HIV viral loads stratified by anti-HBs status.** Median and interquartile ranges are indicated, and there was no significant difference between anti-HBs positive and negative women for either CD4 count or HIV viral loads.

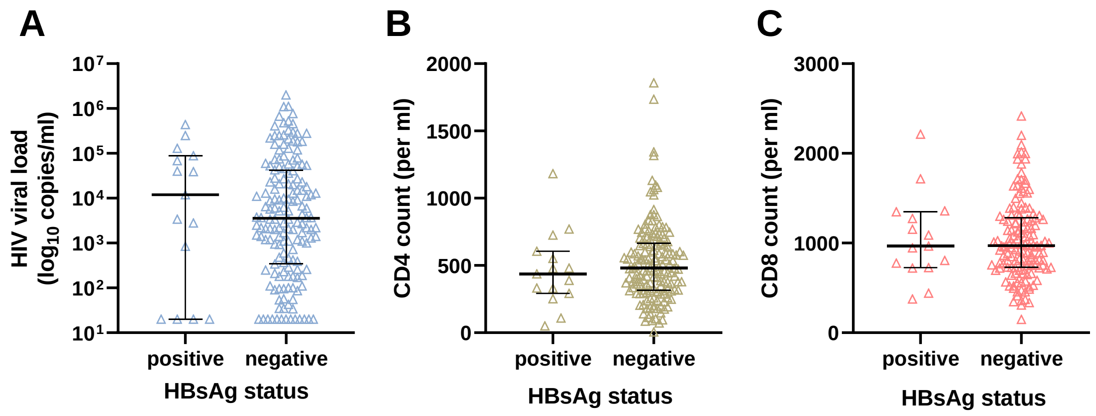


**Supplementary Figure 4; (A) HIV viral load, (B) CD4 count and (C) CD8 count of mothers stratified by HBsAg status.** Median and interquartile ranges are indicated, and there was no significant difference between HBsAg positive and negative women for any biomarkers.

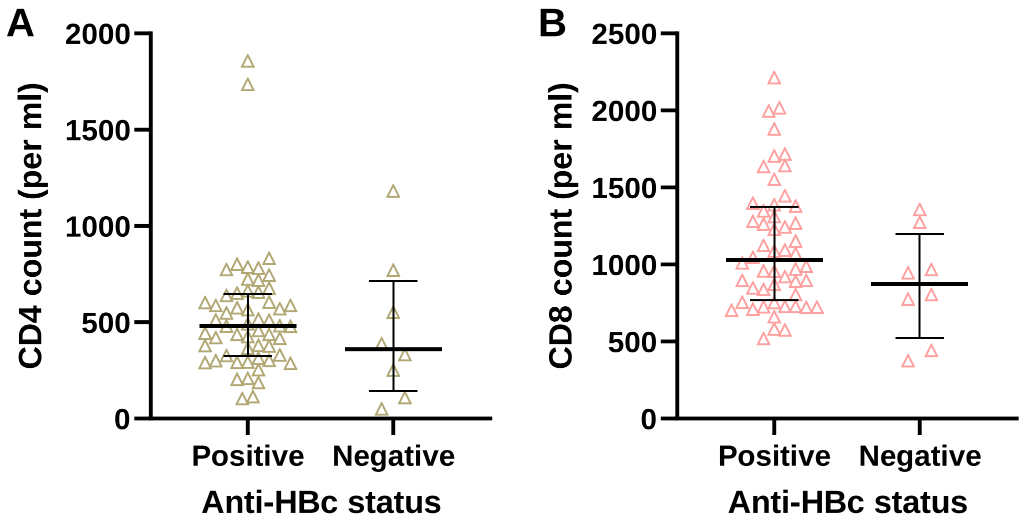


**Supplementary Figure 5; (A) CD4 counts and (B) CD8 counts stratified by anti-HBc status.** All anti-HBc positive women were compared with HBsAg-positive/anti-HBc negative women. Median and interquartile ranges are indicated, and there was no significant difference between anti-HBc positive and negative women for either CD4 or CD8 count.
